# iSCAN: An RT-LAMP-coupled CRISPR-Cas12 module for rapid, sensitive detection of SARS-CoV-2

**DOI:** 10.1101/2020.06.02.20117739

**Authors:** Zahir Ali, Rashid Aman, Ahmed Mahas, Gundra Sivakrishna Rao, Muhammad Tehseen, Tin Marsic, Rahul Salunke, Amit K. Subudhi, Sharif M. Hala, Samir M. Hamdan, Arnab Pain, Norhan Hassan, Magdy M. Mahfouz

## Abstract

The COVID-19 pandemic caused by SARS-CoV-2 affects all aspects of human life. Detection platforms that are efficient, rapid, accurate, specific, sensitive, and user friendly are urgently needed to manage and control the spread of SARS-CoV-2. RT-qPCR based methods are the gold standard for SARS-CoV-2 detection. However, these methods require trained personnel, sophisticated infrastructure, and a long turnaround time, thereby limiting their usefulness. Reverse transcription-loop-mediated isothermal amplification (RT-LAMP), a one-step nucleic acid amplification method conducted at a single temperature, has been used for colorimetric virus detection. CRISPR-Cas12 and CRISPR-Cas13 systems, which possess collateral activity against ssDNA and RNA, respectively, have also been harnessed for virus detection. Here, we built an efficient, rapid, specific, sensitive, user-friendly SARS-CoV-2 detection module that combines the robust virus amplification of RT-LAMP with the specific detection ability of SARS-CoV-2 by CRISPR-Cas12. Furthermore, we combined the RT-LAMP-CRISPR-Cas12 module with lateral flow cells to enable highly efficient point-of-care SARS-CoV-2 detection. Our iSCAN SARS-CoV-2 detection module, which exhibits the critical features of a robust molecular diagnostic device, should facilitate the effective management and control of COVID-19.

## Introduction

In late 2019, an outbreak of the novel *Severe acute respiratory syndrome-related coronavirus* SARS-CoV-2, the causative agent of COVID-19, began in Wuhan, China and rapidly spread throughout the world [1-3]. Based on the data shared by WHO, as of May 24^th^, over five million people have tested positive for COVID-19, with a mortality of over 300,000 people worldwide. Coronaviruses are large, positive-stranded RNA viruses with genome sizes ranging from ~27 to ~32 kb [4]. These viruses pose a serious challenge to human health due to their ability to infect a wide variety of organisms, including avian and mammalian species. The rapid evolution of their genomic RNA by recombination can lead to the generation of viral strains that are more virulent or recalcitrant to therapeutic interventions than the original strains [5]. This is exemplified by the recent emergence and outbreak of the novel SARS-CoV-2.

To curb the spread of this virus, countries throughout the globe have implemented strict measures such as curfews, obligatory quarantines, social distancing, and travel bans. Although these measures have been useful in preventing significant increases in the number of new cases, they have put considerable pressure on the world economy and healthcare systems. Early detection and large-scale screening are crucial for combating emerging infectious diseases, particularly those with symptomatic features that are characteristic of SARS-CoV-2 infection. The facile transmissibility and delay in clinical manifestation of this virus often make it impossible for governments and healthcare providers to assess the gravity of the situation and implement preventive measures to stop the spread of the virus in a timely manner [10]. Unfortunately, as seen in too many cases, a delay in taking decisive action leads to rapid propagation of the virus within a population, where it circulates before suddenly taking on epidemic proportions [11]. From an epidemiological perspective, it is vital to rapidly identify and isolate infected individuals. Timely implementation of counter-measures significantly reduces the burden of the pandemic on society from both a public health and economic standpoint. Furthermore, the lack of testing on an appropriately large scale makes it challenging to keep track of the outbreak and to evaluate the success of the response measures. Therefore, early virus detection and isolation of COVID-19 ‘positive’ individuals are crucial to limit virus transmission and implementation of effective measures to control the spread of the virus and relieve the burden on the healthcare system [12, 13].

Nucleic acid-based diagnostic methods are quite useful and informative compared to serological methods, which are feasible only after antibodies have been produced and only provide information about prior infection and not the current presence of the virus in a patient [14, 15]. The testing and isolation of asymptomatic carriers have proven quite effective in controlling viral spread. Therefore, efficient, low-cost testing methods are required to enable the extensive and recurrent testing of patients. Reverse-transcription PCR (RT-PCR) is the most widely used method for detecting RNA viruses. Due to its sensitivity and specificity, quantitative RT-PCR (RT-qPCR) is currently the gold standard method for identifying the presence of SARS-CoV-2 [1, 16]. Although this method is sensitive and reliable, is not without limitations. RT-qPCR requires highly trained personnel, sophisticated infrastructure, and the transport of samples to central laboratories, thereby limiting its usefulness for large-scale point-of-care diagnostics [17]. In addition, this technique is difficult to implement during emergency situations such as the SARS-CoV-2 pandemic, when thousands of samples must be analyzed as quickly as possible to assess treatment options and prevent further spread of the disease [18]. Therefore, methods that conform to the ASSURED (Affordable, Sensitive, Specific, User-friendly, Rapid and robust, Equipment-free and Deliverable to end-users) features of the point-of-care (POC) diagnostic are urgently needed to mitigate the spread of SARS-CoV-2 through effective control and management measures [19].

Several approaches have been employed for pathogen diagnostics using inexpensive reagents and simple equipment, leading to effective testing in low-resource settings. For example, isothermal amplification approaches based on recombinase polymerase amplification or loop-mediated isothermal amplification (LAMP), which are conducted at a single temperature, have been employed for POC diagnostics for various pathogens, thus providing a very effective alternative to PCR-based methods [14, 20]. Reverse-transcription LAMP (RT-LAMP), which uses a suitable isothermal reverse transcriptase to reverse transcribe a specific sequence of virus RNA to DNA coupled with isothermal reactions such as LAMP, can be used to detect RNA viruses in a single reaction at a single temperature [21]. However, these methods have some drawbacks, including low specificity, a high rate of false positives, and difficulty of adapting these techniques for effective POC diagnostics for pandemics such as COVID-19 [22, 23].

The CRISPR-Cas systems are molecular immunity machines that provide bacterial and archaeal species with immunity against invading nucleic acids, including phages and conjugative plasmids [24, 25]. These systems have been used to engineer the genomes and transcriptomes of all transformable species [26-28]. One intriguing feature of some of these systems, including those employing variants of Cas12, Cas13, and Cas14, is their collateral, non-specific activity following recognition and cleavage of the specific target [29-31]. CRISPR-Cas systems, including CRISPR-Cas12 and CRISPR-Cas13, exhibit robust collateral activity against single-stranded DNA (ssDNA) and RNA targets, respectively. Such collateral activity provides the basis for highly specific, sensitive approaches for nucleic acid detection [32, 33]. For example, SHERLOCK (Specific High-sensitivity Enzymatic Reporter UnLOCKing) and DETECTR (DNA Endonuclease-Targeted CRISPR Trans Reporter) detection have attracted substantial attention [30, 34]. CRISPR-based diagnostic methods exploit the efficiency and simplicity of different isothermal amplification approaches, such as LAMP and recombinase polymerase amplification, which achieve highly specific, sensitive amplification of a few copies of the targeted nucleic acid at single temperature in a short period of time, eliminating the need for thermocycling steps, and are therefore favored for POC diagnostics where low-cost and ease of use are needed. Although these systems hold great promise for developing an effective diagnostic, employing these techniques as POC diagnostics amenable to massive-scale field deployment remains challenging.

In the current study, to address the challenge of SARS-CoV-2 detection, we established a system involving RT-LAMP coupled with CRISPR-Cas12 for the rapid, specific, accurate, sensitive detection of SARS-CoV-2 and named this system iSCAN (***i****n vitro* **S**pecific **C**RISPR-based **A**ssay for **N**ucleic acids detection). Our iSCAN platform is 1) rapid, as the RT-LAMP and CRISPR-Cas12 reaction takes less than 1 h; 2) specific, because detection depends on the identification and subsequent cleavage of SARS-CoV-2 genomic sequences by the Cas12 enzyme; 3) field-deployable, as only simple equipment is required; and 4) easy to use, as the colorimetric reaction coupled to lateral flow immunochromatography makes the assay results easy to assess. Our iSCAN approach is suitable for large-scale, in-field deployment for the early detection of SARS-CoV-2 carriers, allowing them to be effectively isolated and quarantined, thus limiting the spread of the virus. Our SARS-CoV-2 detection kit has been validated using extracted RNAs from clinical samples from COVID-19 positive patients, providing the possibility of widespread deployment for virus detection.

## Results

### Establishment of RT-LAMP for rapid detection of SARS-CoV-2 in a low-resource environment

SARS-CoV-2 testing faces many hurdles, including the availability of critical reagents, highly-trained technical personnel, and sophisticated equipment. Hence, the ability to produce diagnostic reagents in-house is vital in epidemic situations. Therefore, we set out to build a SARS-CoV-2 end-to-end detection platform that is sensitive, specific, easy-to-use, and low cost to facilitate its field deployment on a massive scale. LAMP reactions employ DNA polymerases that possess strand-displacement activities, such as the Bst DNA polymerase large fragment, for efficient amplification of the target DNA [21]. By supplementing the LAMP reaction with a suitable isothermal reverse transcriptase, reverse transcription LAMP (RT-LAMP) can be used to detect RNA by reverse transcribing the target RNA and subsequent DNA amplification in one step. Therefore, we expressed and purified recombinant Bst DNA polymerase large fragment (referred to as Bst LF) from *Geobacillus stearothermophilus* [35], and the synthetic, directed evolution-derived RT ‘xenopolymerase’ RTx enzymes [36]. The catalytic activities and performance of the purified proteins for LAMP with the purified Bst LF, and RT-LAMP with RTx and Bst LF were assayed and compared with commercial Bst and RTx enzymes using published RT-LAMP primers designed against the SARS-CoV-2 genome [37]. Our in-house produced proteins showed efficient and consistent LAMP and RT-LAMP amplifications comparable to the commercial enzymes (Supplementary Figure 1A).

Next, we designed, built, and tested several sets of LAMP primers targeting the SARS-CoV-2 genome. The SARS-CoV-2 genome consists of ~30 kb positive single-stranded RNA with a 5′-cap structure and 3′ poly-A tail containing several genes characteristic of coronaviruses, such as *S* (spike), *E* (envelope), *M* (membrane), and *N* (nucleocapsid) genes (Figure 1A). Other elements of the genome, such as *ORF1a* and *ORF1b*, encode non-structural proteins, including RNA-dependent RNA polymerase (RdRp) [8, 9]. We targeted two regions in the *N* and *E* genes. The *N* gene at the 3′ end of the virus genome is highly conserved among coronaviruses. We designed LAMP primers to generate ~200 bp amplification products to ensure robust amplification sufficient for LAMP-based detection. We conducted RT-LAMP experiments on synthetic SARS-CoV-2 sequences. We identified primer sets, for *N* and *E* genes, which were able to specifically and efficiently amplify the synthetic virus fragments, but not the controls.

**Figure 1:**
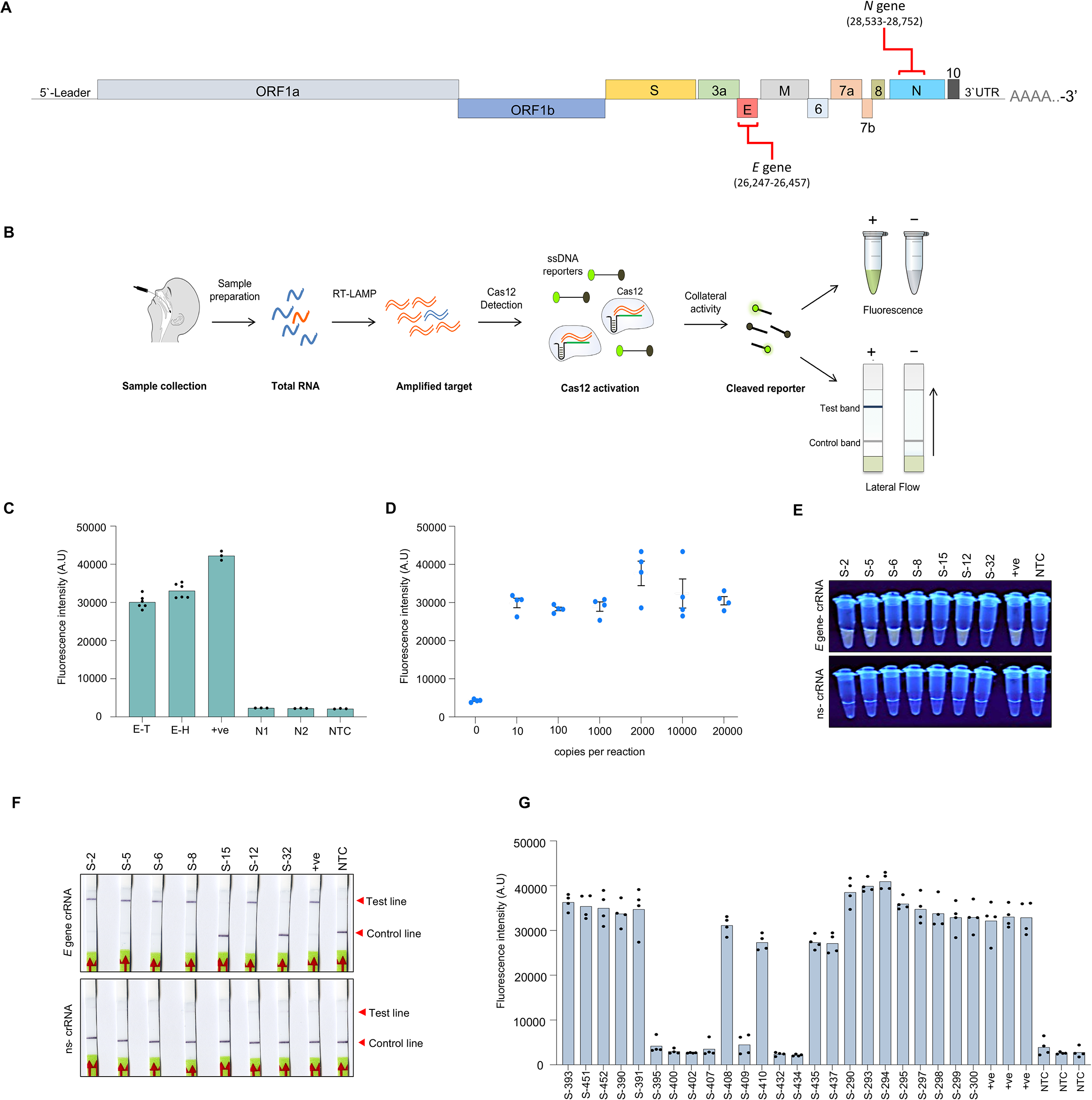
CRISPR–Cas12a-based iSCAN assay for detection of SARS-CoV-2. A- A schematic view of the SARS-CoV-2 genome architecture. Regions targeted by iSCAN assay are highlighted. B- Workflow of iSCAN detection assay. C- Quantifications of signal intensities of CRISPR-Cas12a fluorescence-based detection assays of synthetic SARS-CoV-2. E-T: *E* gene with Tris buffer, E-H: *E* gene with HEPES buffer, +ve: *E* gene with commercial buffer (NEB), N1: no crRNA, N2: ns-crRNA, NTC: no-template control. D- Limit of detection (LoD) determination of RT-LAMP-Cas12a assay. Values shown as mean ± SEM (n=4) E- End-point fluorescence visualization under UV light following the CRISPR/Cas12a detection assay performed on clinical samples. S: different clinical samples with different Ct values, +ve: Synthetic RNA, NTC: no-template control. F- Lateral flow readouts of Cas12a detection of SARS-CoV-2 RNA in clinical samples G- End-point fluorescence intensity measured in clinical samples after CRISPR/Cas12a detection assay.

### Efficient detection of SARS-CoV-2 via RT-LAMP coupled with CRISPR-Cas12a

The sensitivity of RT-LAMP is comparable with the RT-PCR. However, the specificity of the reactions and the visualization of the results may be complicated due to primer-dimer formation, non-specific amplification, and cross-contamination when running a large number of samples simultaneously. Therefore, to develop a highly sensitive and specific detection system, we coupled CRISPR-Cas12 with RT-LAMP target amplification to establish a binary system for SARS-CoV-2 detection. To this end, we purified the class II, type V *Lachnospiraceae bacterium* ND2006 (LbCas12a) orthologue.

We assessed its programmable (specific) *cis*-cleavage activity against dsDNA targets and (collateral) *trans*-cleavage activity on ssDNA non-targeted sequences. Purified LbCas12a assembled with crRNAs targeting dsDNA fragments of the SARS-CoV-2 *N* or *E* genes exhibited endonuclease activity on the targeted dsDNA, confirming the catalytic *cis*-cleavage activity of the purified Cas12a (Supplementary Figure 1B). To assess the collateral activity of the purified LbCas12a protein, the specific dsDNA targeting reaction was supplemented with non-targeted fluorophore quencher (FQ)-labeled ssDNA reporters. Cas12a catalyzed the efficient degradation of the ssDNA FQ reporters as measured by the fluorescent signal, which was generated only in the presence of the specific dsDNA targets and specific crRNA, indicating the active collateral cleavage activity of the purified enzyme (Supplementary Figure 1C, D).

To establish our iSCAN system for the detection of SARS-CoV-2, we used RT-LAMP for virus detection and amplification coupled with CRISPR-Cas12 as a specificity factor. The positive signal from Cas12 provides a specific and accurate signal for virus detection. Collateral ssDNA-FQ reporter is cleaved upon Cas12a binding and cleavage of the target virus sequence in the RT-LAMP amplification products (Figure 1B). We designed crRNAs specific to the SARS-CoV-2 *N* and *E* gene genomic regions amplified with the primer sets identified in the primer screening assays.

We tested the activity and specificity of our two-pot iSCAN system using an FQ reporter assay. We performed RT-LAMP using the *E* gene-specific RT-LAMP primers and synthetic SARS-CoV-2 genome. Subsequently, we introduced the RT-LAMP product to Cas12a protein pre-assembled with crRNAs specific to the *E* gene amplicon, non-specific crRNA, or without crRNA in the presence of FQ-ssDNA reporter. Cas12a cleaved the ssDNA reporters only in the presence of specific crRNA and RT-LAMP amplicons, confirming the specificity and activity of our systems (Figure 1C).

Next, we quantitatively compared the sensitivity of our system to that of the approved SARS-CoV-2 CDC RT-qPCR assays to determine the limit of detection (LoD). We conducted our two-pot iSCAN assays with various dilutions of the synthetic viral RNA ranging from 0 to 20000 copies per reaction. The LoD assays showed that the iSCAN reactions could detect down to 10 RNA copies per reaction, indicating the high sensitivity of our assay (Figure 1D).

To further optimize the assay, we systemically evaluated different RT-LAMP and Cas12a detection timing to determine the time needed to detect the signal. Using the lowest concentration identified in the LoD assay, we added RT-LAMP products from different amplification time points, including 5, 10, 20, and 30 min to crRNA-Cas12a detection complex along with the fluorescence reporter. The samples were incubated at 37°C for different times (5, 10, 20, and 30 min), and the fluorescent signal generated from the reporter by the Cas12a collateral activity upon target recognition was measured. Our assay reliably detected the positive signal with 20 min of RT-LAMP followed by 20 min of Cas12a detection comparable to the stronger fluorescent signal with prolonged RT-LAMP and Cas12a detection (Supplementary Figure 2).

### Validation of RT-LAMP coupled CRISPR-Cas12a for the detection of SARS-CoV-2 in clinical samples

To validate our detection system for application with real samples from patients, we first assessed the capability of our iSCAN assay to detect SARS-CoV-2 nucleic acid extracted from nasopharyngeal swabs of five different patients who tested positive for SARS-CoV-2 with RT-qPCR assays and two patients who tested negative (Supplementary File 2). The positive samples had different Ct values ranging from 15 to 40 with the CDC qPCR *N* gene primer sets (IDT, Catalogue #10006606). Using fluorescence-based detection with at least three replicates for each sample, our iSCAN system targeting the SARS-CoV-2 *E* gene showed 100% agreement with RT-qPCR results (Figure 1E and Supplementary Figure 3).

To facilitate the effective detection of SARS-CoV-2 in a low-resource environment, we sought to simplify the use of iSCAN for the detection of SARS-CoV-2. Therefore, we coupled lateral flow immunochromatography with our iSCAN detection system. Lateral flow strips are user-friendly and straight-forward, and can facilitate the development of home testing for SARS-CoV-2. We evaluated our system for POC diagnostics with lateral flow readouts using commercial lateral flow strips. Lateral flow assays showed 100% concordance with results obtained with fluorescence-based detection (Figure 1F).

Next, we evaluated our iSCAN assay by testing on total RNAs extracted from an additional 24 nasopharyngeal swab samples obtained from 21 SARS-CoV-2-positive and 3 negative patients (Supplementary File 2). We assayed the performance of our system with *E* and *N* gene primers and crRNAs. We observed low sensitivity of our assay when using the *E* gene target, with 8 out of 21 positive samples testing positive, and 3 out of 3 negative samples testing negative with fluorescence-based readouts (Supplementary Figure 4). However, when we used the *N* gene assay, we observed a significant increase in the sensitivity of the assay, where 18 samples tested positive out of the 21 positive samples with qPCR (~86%), and the 3 negative samples were diagnosed correctly in agreement with the qPCR data (100%) (Figure 1G). We next tested the performance our iSCAN *N* gene assay using the simple lateral flow readouts. We observed high agreement between the fluorescent-based detection and the lateral flow readout results (Supplementary Figure 5).

### SARS-CoV-2 detection via RT-LAMP coupled CRISPR-Cas12b

Our two-pot iSCAN systems, employing RT-LAMP coupled with CRISPR-Cas12a, reliably and specifically detected SARS-CoV-2 from clinical patient samples. However, developing virus detection modalities suitable for POC testing might require minimal liquid handling and single-pot reactions to facilitate wide adoption and in-field deployment. Therefore, we attempted to further simplify our iSCAN system and develop a one-pot assay by employing the thermophilic variants of Cas12b that can catalytically function in the same temperature range as RT-LAMP. To this end, we purified Cas12b from *Alicyclobacillus acidoterrestris* (AacCas12b) and Cas12b from *Alicyclobacillus acidophilus* (AapCas12b) variants [38, 39]. We confirmed the *cis* activity of these proteins on PCR products of the target sequences using sgRNA sequence of the AacCas12b variant. Our data show that both Cas12b variants were capable of inducing double-strand breaks using the same sgRNA at 62°C, and had robust collateral activities when incubated with ssDNA FQ reporter (Supplementary Figure 6). However, because AapCas12b showed enhanced *cis* cleavage activity at 62°C, we chose to proceed with this Cas12b variant.

The goal of utilizing the thermophilic Cas12b variants was to develop a simple one-pot and single temperature detection modality that minimizes liquid handling. This can be achieved by mixing all amplification and CRISPR-based detection reagents simultaneously in one pot, which allows simultaneous target amplification and detection. Thus, using the *N* gene assay, we attempted to verify the feasibility of performing SARS-CoV-2 detection in a one-pot assay format using the synthetic virus RNA in one-pot detection reaction mixtures containing AapCas12b, sgRNA, ssDNA-reporters, and RT-LAMP amplification reagents, and incubated the reaction mixtures at 62°C for 1 h. The one-pot detection system was capable of detecting the viral RNA, albeit at lower efficiency compared to the two-pot system (Supplementary Figure 7A). When we visualized the RT-LAMP product from one-pot assay on an agarose gel, we found weak amplification of the target virus RNA. Therefore, we hypothesized that the concurrent presence of the active Cas12b–sgRNA complex in the same pot leads to the digestion of the initial RT-LAMP product, which significantly affects the performance of the RT-LAMP amplification, and thus the robustness of the detection.

To improve the performance of the one-pot detection, we separated the Cas12b enzyme from the rest of the detection components by adding the Cas12b protein in a droplet on the tube wall, then allowed the RT-LAMP reaction to proceed before mixing the Cas12b with the other reaction components. We observed an enhanced RT-LAMP amplification of the target and improved detection performance using this “spotted” one-pot reaction with synthetic viral RNA (Supplementary Figure 7A). In the spotted one-pot reaction, AapCas12b consistently achieved a LoD of 10 copies per reaction (Figure 2A and Supplementary Figure 7B).

**Figure 2:**
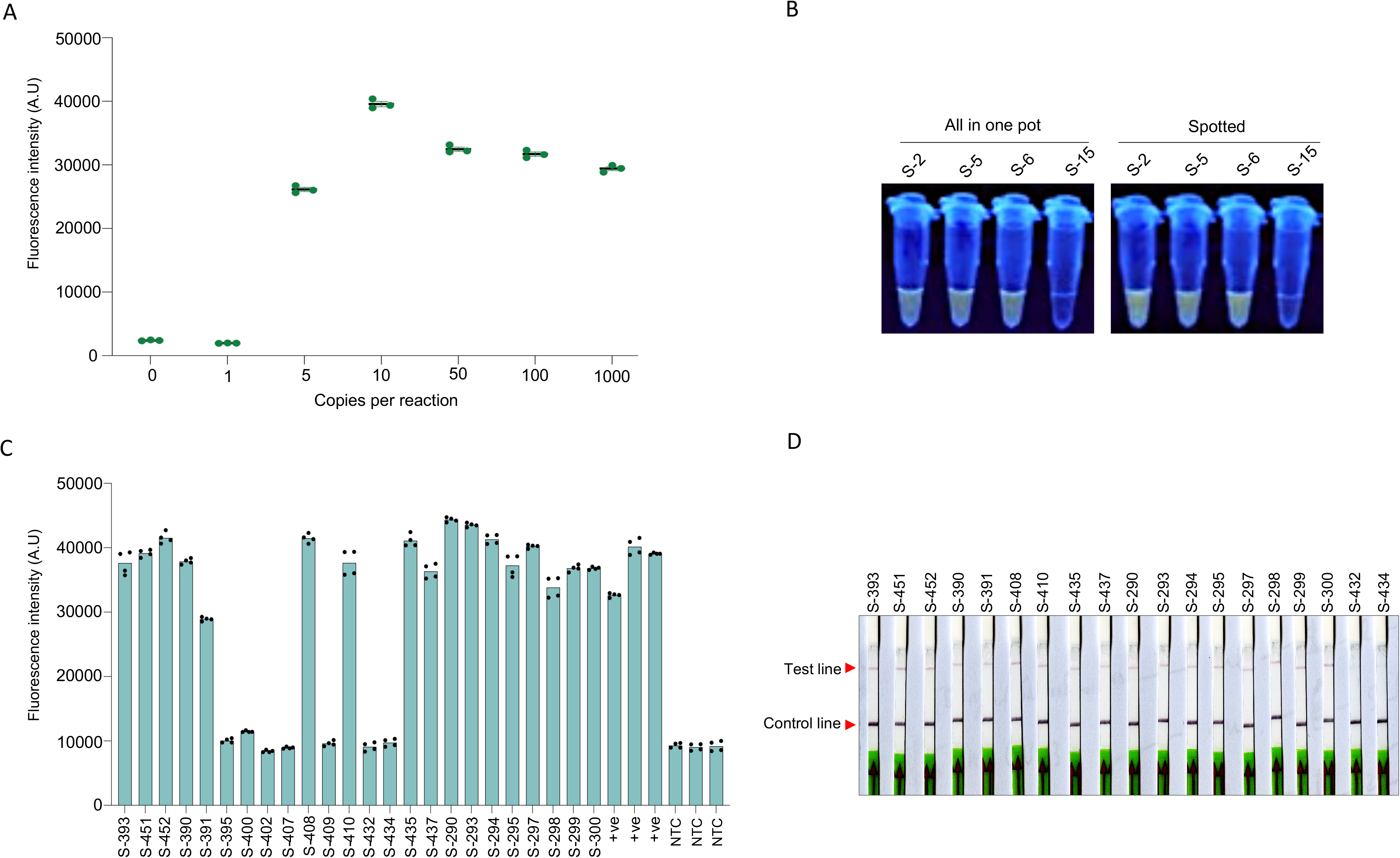
CRISPR–Cas12b-based assay for detection of SARS-CoV-2. A- Limit of detection (LoD) of RT-LAMP-AapCas12b assay in spotted one-pot reaction. Values shown as mean ± SEM (n= 3). B- Visual fluorescence output of four clinical samples under UV light following CRISPR/Cas12b detection assay performed by two different strategies. S: different clinical samples with different Ct values. C- End-point fluorescence intensity measured in clinical samples after performing spotted one-pot CRISPR-AapCas12a detection assay. S: different clinical samples with different Ct values, +Ve: Synthetic RNA, NTC: no-template control. D- Lateral flow readout after spotted one-pot AapCas12b detection assay performed on clinical samples.

Next, we tested the ability of the AapCas12b-based detection system to detect SARS-CoV-2 RNAs in clinical samples (3 positive and 1 negative) using fluorescence-based and lateral flow-based readouts. AapCas12b was capable of detecting the SARS-CoV-2 RNA using spotted one-pot and all-mixed one-pot reactions (Figure 2B and Supplementary Figure 7C). Using the spotted one-pot AapCas12b detection system, we tested the 24 clinical samples used with the Cas12a based detection system. The performance of AapCas12b system was very similar to the performance of Cas12a-based system, where 18 out of the 21 qPCR-positive samples showed a positive signal, and the 3 qPCR negative samples showed negative results using fluorescence-based readouts (Figure 2C). However, when we tested the system on 17 of the positive samples and 2 negative samples using lateral flow readouts, we consistently observed weak performance measured by the weak signal on lateral flow readouts in positive samples compared to negative ones, which compromised the interpretation of the results (Figure 2D).

## Discussion

The COVID-19 outbreak poses an unprecedented public health challenge worldwide. The crucial need for the large-scale detection of SARS-CoV-2 has made it urgent to develop local solutions for creating sensitive, specific, low-cost, in-field deployable diagnostic kits. The bio-manufacturing of assay reagents and simple portable machines for easy use at POC will facilitate the widespread, large-scale detection of this virus. In this study, we established iSCAN, a system involving RT-LAMP coupled with CRISPR-Cas12, as an efficient detection module for COVID-19. We designed, screened, and tested different sets of LAMP primers and identified a few primer sets for specifically and efficiently amplifying the SARS-CoV-2 *E* and *N* genes. Due to huge disruptions in the supply chain worldwide, we employed the in-house purified RTx and BstI enzymes for the RT-LAMP reactions.

The addition of CRISPR reagents provides important specificity to the assay. Although the results of LAMP reactions can be visualized with a pH-sensitive dye upon DNA amplification, false amplification, cross-contamination, and the formation of primer-dimers may lead to false signals [22, 23]. Therefore, a specificity factor is needed to ascertain the identity of the amplification products and whether these products are derived from virus sequences. Because CRISPR-Cas12 cuts DNA only in the presence of the SARS-CoV-2 sequence, the Cas12 enzyme serves as a specificity factor for RT-LAMP. RT-LAMP polymerases can be used for reverse transcription of the viral genome and subsequent amplification, producing a sufficient DNA template for CRISPR-Cas12 to enable complete virus detection in 1 h in one-pot or two-pot assays, depending on the Cas12 variant employed in the detection system. We optimized RT-LAMP to successfully and reproducibly detect SARS-CoV-2. To ensure that the amplified RT-LAMP products are derived from genuine virus sequences, we coupled RT-LAMP and CRISPR-Cas12-based detection. RT-LAMP coupled with CRISPR-Cas12 offers a binary result of RT-LAMP and Cas12 activity, providing a highly specific, robust virus detection modality for SARS-CoV-2.

The LoD of the iSCAN system is comparable with the minimum level required to detect the virus in clinical samples by qRT-PCR. Minimal sample processing is sufficient for the iSCAN system to detect SARS-CoV-2, enabling large-scale screening. Our iSCAN system for SARS-CoV-2 detection is portable and can easily be deployed in the field due to its simplicity and low cost (2–5 USD/reaction).

We have adjusted our iSCAN assays to make them amenable to use by lay users without specialized equipment (Figure 3), but have retained some complexity to assure accurate assays. For example, although one-pot detection systems are favored over two-pot detection systems for onsite detection (Figure 3), our data show that mixing various components in one pot leads to a substantial reduction in detection performance and sensitivity. Further optimization of the one-pot detection assay could help improve the performance of this system. Such optimization would involve screening different Cas12b variants and testing different isothermal amplification methods. Cas12a can be further developed as a one-pot detection system by employing special tubes that allow some reaction components to be isolated before use.

**Figure 3:**
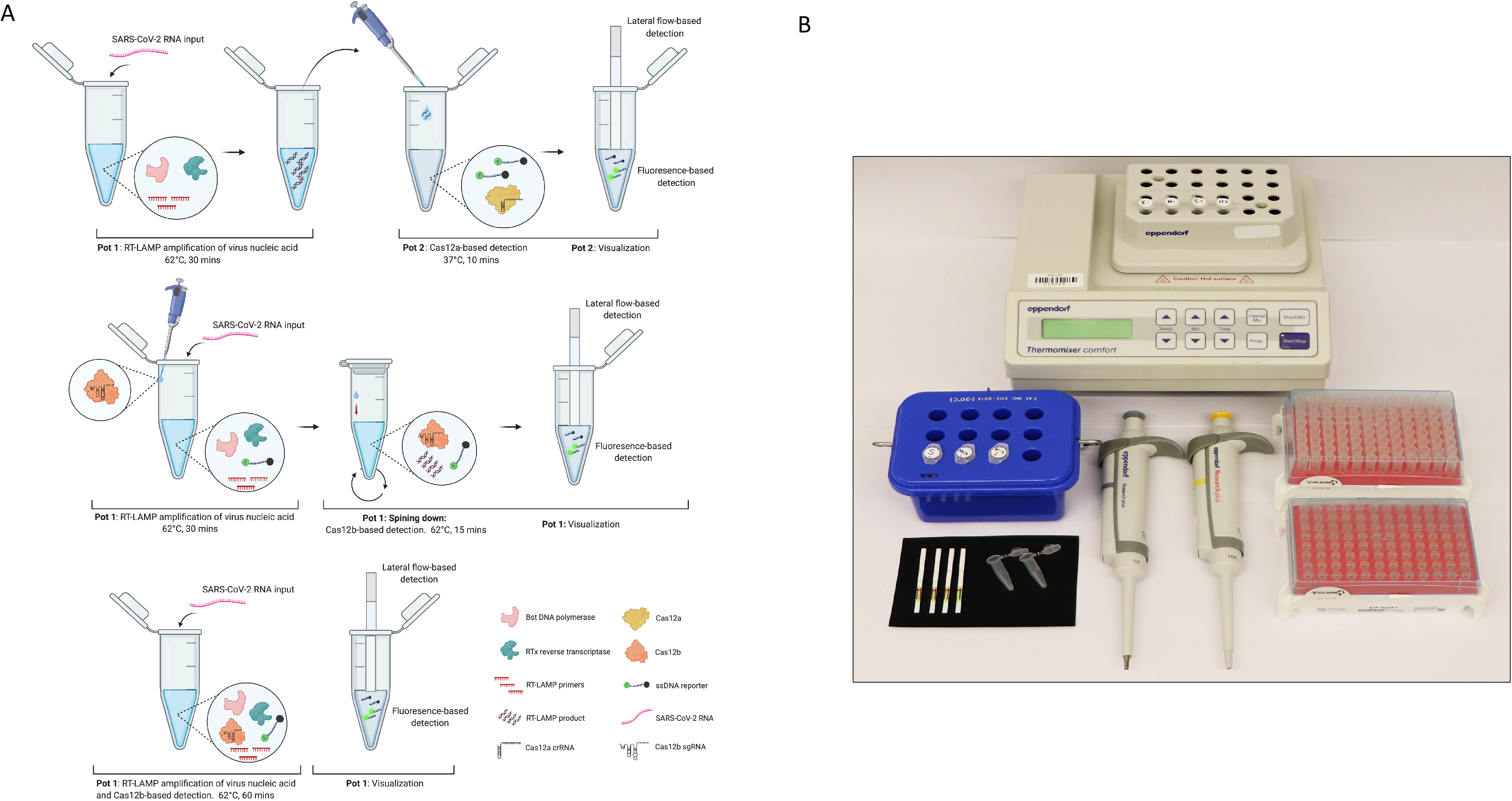
Overview of the CRISPR-Cas based SARS-CoV-2 detection methods and requirements. A) Schematic illustration showing different Cas12-based detection modalities. The first row depicts Cas12a-based two-pot reaction, where the RT-LAMP amplification of SARS-CoV-2 nucleic acid is performed in the first tube. Then the RT-LAMP product is transferred to a second tube for Cas12a-based detection. The second row depicts RT-LAMP and Cas12b-based detection reagents, except for Cas12b-sgRNA complex, mixed in a single reaction. Cas12b-sgRNA complex is temporary separated from the reaction because it is added on the tube wall. After 30 min of RT-LAMP reaction, the Cas12b-sgRNA complex is centrifuged into the reaction mix for target cleavage and CRISPR/Cas-based detection, which takes place for an additional 15 min. In the third row, simultaneous RT-LAMP amplification and CRISPR-based detection of SARS-CoV-2 detection is performed in a single tube. B) Minimal equipment and consumables required for iSCAN detection assay, which includes a heat block, pipettes, pipette tips, sample tubes, and lateral flow strips.

The iSCAN system can be further simplified through the use of cellular extracts harboring assay reagents to maximize product integrity during shipping and handling. This would allow a lay user to simply mix, incubate, and vortex the sample and visualize the assay results. The large-scale production of these reagents in various forms is feasible, including bacterial extracts or freeze-dried cells, allowing large-scale in-field deployment in low resource environments [40]. The calorimetric visualization of the test results does not require special equipment and can be conducted by the naked eye or using a simple portable machine. We also predict that manifolds for both the RT-LAMP and CRISPR-Cas12 reactions could be designed to enable sample processing, reaction mixing, and assay results to be obtained in a closed system to allow non-healthcare staff to perform these assays.

In addition to providing high specificity, the iSCAN assays can be easily customized as the virus mutates. The iSCAN two-pot and one-pot systems using RT-LAMP coupled with CRISPR-Cas12 allow SARS-CoV-2 to be visually detected with a detection limit comparable to that of qRT-PCR. This detection module couples the robustness of the RT-LAMP assay with the specificity of CRISPR-Cas12, resulting in an efficient and reproducible detection system. Because new mutations could occur in the current virus strain, iSCAN can easily be tailored to detect any virus strain and to differentiate between different strains and determine the dominance of a specific strain in a certain location or population.

Moreover, the iSCAN assays can be used for high-throughput testing. Various detection modules, including GeneXpert, Cephied, and ID NOW by Abbot are low throughput and require the use of a single machine for each sample, which may be impractical for low-resource environments and where large-scale testing in a short time window is needed. Because asymptomatic virus transmission has been reported, and initially negative results may not be conclusive, regular testing may be required for patients with symptoms and their asymptomatic primary and secondary contacts. Our iSCAN system is suitable for conducting multiple consecutive tests at the POC. Because the viral load in a patient plays a significant role in the assay results, regular testing is required. The development of the iSCAN system as a POC SARS-CoV-2 testing modality would enable affordable and regular testing, which is crucial for reducing false negatives and identifying clusters of infection as early as they arise. To facilitate sample collection and testing, a patient’s saliva could be used for virus detection [41], and the iSCAN module could be developed as an in-home SARS-CoV-2 testing kit in the future. Our iSCAN detection platform can be designed to differentiate between coronaviruses and identify specific SARS-CoV-2 strains. Therefore, the iSCAN detection system could greatly facilitate the effective management and control of virus spread, specifically in high-risk areas, including clinics, airports, and low resource areas where the test could be conducted and results obtained in a short time.

The iSCAN detection module might not require a nucleic acid extraction step and may be compatible with different types of samples, including saliva and nasal swabs. Furthermore, quick nucleic acid extraction could be coupled with iSCAN to speed up the detection and bypass the need to conduct an RNA extraction step. We envision that iSCAN could be used to assess patient-collected saliva samples in a tube with extraction buffer that is compatible with RT-LAMP and CRISPR-Cas12 reactions. The iSCAN detection module will ensure the safety of both healthcare personnel and laboratory staff. We aim to develop the current iSCAN system further for lay users and POC as well as in-home testing, which will help produce quick results and prevent transmission of the virus. Therefore, our iSCAN system could help provide physicians and healthcare staff with the answers they need for better, more timely patient-management decisions to mitigate the spread of the virus and its impact on the overwhelmed healthcare system during the outbreak.

Other versions of iSCAN could be developed in the future for multiplex targeting of different virus strains and the simultaneous identification of other pathogens. Although we have shown that iSCAN works quite efficiently with lateral flow cells, inexpensive visualizers of the reporters could be employed to increase the throughput of the assays and more rapidly produce results from a large number of samples. Furthermore, this represents an effective alternative should the supply of lateral flow cells become limited or disrupted. Other DIY modalities could be developed to couple the assay reactions and test results with a smartphone to allow the results to be communicated rapidly. In conclusion, the iSCAN virus detection platform should enable robust testing on a massive scale in low-resource environments, thereby helping to mitigate the transmission and spread of SARS-CoV-2 and providing effective management of the COVID-19 pandemic.

## Material and methods

### Nucleic acid preparations

The control SARS-CoV-2 viral RNA sequences used in this study were synthetic RNA from Twist Bioscience, 102024 diluted to (1×103 RNA copies/microliter). For Cas12 DNA cleavage assays, dsDNA substrates were generated by either PCR amplification from SARS-CoV2_N_Positive Control vector (IDT, 10006625) using forward (N gene-F) and reverse primer (N gene-R) (Supplementary Table 2) for *N* gene, or was designed and ordered as dsDNA gBlocks (IDT) for *E* gene (Supplementary Table 3). The gblocks were cloned into pJet2.1 (Thermo Scientific, K1231) vector and the resulting plasmids harboring the clones were used for PCR. PCR products were gel purified using the QIAquick Gel Extraction kit following manufacturer’s instructions. To assess the activity of the purified Bst LF via LAMP reaction, the *N* gene PCR amplicon was used as template in the LAMP reaction. *In vitro* transcription was carried out using TranscriptAid T7 High Yield Transcription Kit (Thermo Scientific K0441) following manufacturer’s instructions. For LbCas12a crRNAs, templates for *in vitro* transcription were generated using single-stranded DNA oligos containing a T7 promoter, scaffold and spacer in reverse complement orientation (IDT), which were then annealed to T7 forward primer in taq DNA polymerase buffer (Invitrogen). For Cas12b sgRNAs, the AacCas12b 90 nt-long sgRNA scaffold was synthesized and ordered as sense and anti-sense ssDNA ultramers containing the T7 promoter at the 5′ end (IDT). The two ssDNA strands were annealed in taq DNA polymerase buffer to generate T7-Cas12b sgRNA scaffold. The scaffold was used as a template for PCR with T7 forward primer and reverse primers containing 20 nt spacer sequences. All DNA oligos and substrates are listed in (Supplementary Table 2 and 4) The *in vitro* transcribed sgRNAs were purified using MEGAclear™ Transcription Clean-Up Kit (Thermo Scientific AM1908) following manufacturer’s instructions.

### Protein purification

Bst LF DNA polymerase was induced at OD_600_ 0.7 with 100 ng/mL tetracycline-HCl at 37 °C for 3 hours and the pellet was collected. Bst LF DNA polymerase was purified following *Li et. al* 2017 protocol [35, 42]. The collected protein fractions were quantified, and aliquots were flash freezed in liquid nitrogen. RTx reverse transcriptase was purified following Bhadra *et. al* 2020 protocol [43]. The collected protein fractions were quantified, and aliquots were flash freezed in liquid nitrogen. LbCas12a, AacCas12b, AapCas12b recombinant proteins were purified following Chen *et. al* 2018 protocol [30]

### Assessment of the activity and functionality of in-house produced enzymes

To confirm the polymerization activity of the purified Bst LF DNA polymerase enzyme LAMP reaction was assembled and carried out at 62 °C. The commercial Bst enzyme Bst 2.0 WarmStart DNA Polymerase (NEB) was used as per manufacturer’s instructions as control. As initial confirmation, the reported LAMP primers designed against SARS-CoV-2 were used [37] (Supplementary table 1). The activity of the purified RTx reverse transcriptase enzyme was confirmed by single enzyme RT-PCR reaction following Bhadra *et. al* 2020 protocol [43]. The Synthetic SARS-CoV-2 RNA control (102024 Twist Bioscience) was used as input RNA template. The one step RT-PCR was carried out with cycling conditions: 60°C for 30 minutes, 95°C for 10 minutes and 45 cycles of 95°C for 15 seconds followed by 55°C for 30 seconds and 68 °C. The amplified product was separated on 1% agarose gel stained with SYBR-safe dye. The dsDNA cleavage and collateral ssDNA degradation activities of Cas12a and Cas12b proteins were confirmed following previously described protocols [30, 38]

### Design and screening of LAMP primers

Different primer sets targeting several regions of the N gene and E gene of the SARS-CoV-2 genome (GenBank accession number MN908947) were designed using PrimerExplorer v5 software (https://primerexplorer.jp/e/). Optimal sets that showed the best performance were determined by conducting LAMP assays to detect specific targets. (Supplementary Table 1). Screening of the best performing and most sensitive LAMP primers was done by running RT-LAMP as described below.

### Two-pot iSCAN detection assay

The iSCAN detection assays were performed in two steps. In the first step, RT-LAMP was performed to reverse transcribe and pre-amplify the viral RNA to generate dsDNA substrates for Cas12 enzymes. All the reagents for LAMP reactions were assembled on ice and combined in a single reaction mixture of 2.5 μL 10X isothermal amplification buffer (1X, 20 mM Tris-HCl (pH 8.8), 10 mM (NH_4_)_2_SO_4_, 50 mM KCl, 2 mM MgSO_4_, 0.1% Tween 20), 2.5 μL 10X Primer Mix (2 μM F3 2 μM B3 16 μM FIP 16 μM BIP 8 μM LF 8 μM LB), 1.13 μL of 100 mM MgSO_4_, 1.4 μL of 10mM LF DNA polymerase (50 ng), 1 μL of RTx reverse transciptase 4 ng/μl and nuclease-free water to 25 μL. Mixtures were incubated at 62°C for 30 minutes or as indicated.

Next, for Cas12 detection assays, 250 nM LbCas12a (500 nM for AacCas12b and AapCas12b) was first pre-incubated with 250 nM (500 nM for AacCas12b and AapCas12b) of specific (or non-specific) LbCas21a crRNAs in 1x Cas12 reaction Buffer for 30 min at 37 °C (62 °C for Cas12b) to assemble Cas12-crRNA ribonucleoprotein (RNP) complexes. The reaction was diluted 4 times with 1x binding buffer. For fluorescence-based detection, for Cas12a 250 nM and for Cas12b, 500 nM of fluorophore-quencher (FQ) reporter (5'-/56-FAM/TTATT/3IABKFQ/-3', IDT) or DNase alert substrate (11-02-01-02, IDT) and 2ul of the RT-LAMP reaction was added to the pre-assembled Cas12-sgRNA RNP complexes and incubated at 37 °C (62 °C for Cas12b) for 10 - 30 mins. End-point fluorescence detection was monitored using a Tecan plate reader (Tecan 200).

For lateral flow detection, the Cas12 detection reaction was assembled as described above with modifications. Lateral flow cleavage reporter (5'-/56-FAM/TTATTATT/3Bio/-3', IDT) was added to the reaction at a final concentration of 500 nM in 100 ul reaction volume along with the 2 ul RT-LAMP product, and the reaction was incubated at 37C° (62 °C for Cas12b) for 10 – 30 minutes. Upon completion of the reaction, lateral flow readouts were performed by placing lateral flow dipsticks (Milenia HybriDetect 1, TwistDx, cat. no. MILENIA01) into Cas12 detection reaction and results were observed after 5 minutes. Interpretation of lateral flow results was done as follows, a single band, close to the sample pad indicated a negative result (control band), whereas a single band close to the top of the strip (test band) or two bands (test and control) indicated a positive result (Figure 1B).

### One-pot iSCAN detection assays

To develop one-pot iSCAN detection system with AapCas12b, we employed different strategies; L of 5 μL of 5μM AapCas12b was spotted to the walls of the tube [44, 45] or directly added to the initial 50 μL reaction mix, independently. The initial 50 μL of reaction mix contains the complete RT-LAMP reagents such as 5 μL 10X isothermal amplification buffer, 5 μL 10X Primer Mix, 4 μL of 100 mM MgSO_4_, 7 μL of 10 mM dNTP mix, 2 μL of Bst LF DNA polymerase (50 ng), and 2 ul of RTx reverse transciptase 4 ng/μl, together with 2.5 μL of 500 ng/μL salmon sperm DNA, 2 μL of viral (control or clinical sample) RNA, 5 μL of 5 μM specific (or non-specific) AacCas21b sgRNAs and 5 μL of 5 μM lateral flow cleavage reporter (5'-/56-FAM/TTATTATT/3Bio/-3', IDT) and 5.5 μL nuclease-free water. After the addition of all reagents we incubated the reaction tubes horizontally in a water bath at 62°C for 30 minutes and then spin down the tubes to mix the AapCas12b with the LAMP reaction and incubated the reaction tubes again at 62°C for 30 minutes for further AapCas12b based collateral activity. In case of AapCas12b direct addition to the total reaction mix, we incubated the tubes in heat block at 62°C for 1 hr. Each reaction was carried out in duplicates. Later, for the lateral flow-based detection we combined the two 50 μl duplicate reaction mixes into one tube (100 μl and carried out lateral flow detection as described earlier.

### Clinical sample collection and RNA extraction

Oropharyngeal and nasopharyngeal swabs were collected by the physicians from COVID-19 suspected patients in Ministry of Health hospitals in the Western region in Saudi Arabia and placed in 2-mL screw-capped cryotubes containing 1 mL of TRIZOL for inactivation and transport. The sample tubes were sprayed with 70% ethanol enveloped with absorbent tissue and placed in individual and sealed biohazard bags with labels accordingly. The bags are then placed in leak-proof boxes and sprayed with 70% ethanol before placing them in a dry ice container for subsequent transfer to King Abdullah University of Science and Technology (KAUST). Total RNAs from the samples (that contained the SARS-CoV-2 virus RNAs) was then extracted following instructions as described in the CDC EUA-approved protocol using the DIRECTZOL KIT from ZymoBiomics (Direct-Zol RNA Miniprep (Zymo Research); catalog #R2070) following the manufacturer’s instructions.

### Real-time reverse transcription PCR (RT-PCR) for detection of positive SARS-CoV2 RNA sample

RT-PCR was conducted by using the oligonucleotide primer/probe (Integrated DNA Technologies, catalog #10006606) and Superscript III one-step RT-PCR system with Platinum Taq Polymerase (catalog #12574-026) as per the manufacturer’s protocol on extracted RNA sample. The RNA sample for SARS nCoV 2019 considered as positive when the cycle threshold (Ct) value for both *E* and *N* genes was ≤ 36 and negative when the Ct value was more than > 36 for both genes.

## Data Availability

All data are available in the supplementary information.

## Acknowledgments

We would like to thank members of the genome engineering and synthetic biology laboratory for insightful discussions and technical support. We thank Professor Andrew Ellington for providing the RTx and BstI clones. This work was supported, in part, by the Smart Health Initiative at KAUST and the IAF grant from the KAUST IED to MM. The samples were collected from patients providing verbal consent to the treating doctor after they read the patient consent form, or the doctor explained the content of consent form to the patient. Hence, as per the approved protocol, the doctors took samples only after receiving verbal consent from a concerned patient (due to infection hazard and no family member being available next to the patients due to infection control guidelines) and the treating doctor signed on the consent form on behalf of the patient before any sample collection took place. We have obtained the necessary ethical approval from the Saudi-MOH and the Mass Gathering directorate in Riyadh, Saudi Arabia, for the collection of the samples (Saudi MOH Approval #: H-02-K-076-0420-285, dated 13.04.2020; KAUST Approval #: 20IBEC14). We thank Sara Mfarrej and Fathia Ben Rached for their help during the handling of the clinical samples in KAUST.

## Notes

### Competing Interest Statement

ID has been filed based on the methods used in this paper

### Author Declarations

We have obtained the necessary ethical approvals from the Saudi MOH and the Mass Gathering directorate in Riyadh for the collection of the samples (MOH Approval #: H-02-K-076-0420-285, dated 13.04.2020; KAUST approval #: 20IBEC14) by AP.

